# Effectiveness of Covid-19 Vaccines in the United States Over 9 Months: Surveillance Data from the State of North Carolina

**DOI:** 10.1101/2021.10.25.21265304

**Authors:** Dan-Yu Lin, Yu Gu, Bradford Wheeler, Hayley Young, Shannon Holloway, Shadia Khan Sunny, Zack Moore, Donglin Zeng

**Author notes:** Address correspondence to Dr. Dan-Yu Lin, Department of Biostatistics, University of North Carolina, Chapel Hill, NC 27599-7420, USA.

## Abstract

**Background:** The duration of protection afforded by Covid-19 vaccines in the United States is unclear. Whether the recent increase of breakthrough infections was caused by waning immunity to the primary vaccination or by emergence of new variants that are more highly transmissible is also unknown.

**Methods:** We extracted data on vaccination histories and clinical outcomes (Covid-19, hospitalization, death) for the period from December 13, 2020 through September 8, 2021 by linking data from the North Carolina COVID-19 Surveillance System and COVID-19 Vaccine Management System covering ∼10.6 million residents statewide. We used the Kaplan-Meier method to estimate the effectiveness of the BNT162b2 (Pfizer–BioNTech), mRNA-1273 (Moderna), and Ad26.COV2.S (Janssen) vaccines in reducing the incidence of Covid-19 over successive post-vaccination time periods, producing separate estimates for individuals vaccinated during different calendar periods. In addition, we used Cox regression with time-dependent vaccination status and time-varying hazard ratios to estimate the effectiveness of the three vaccines in reducing the hazard rates or current risks of Covid-19, hospitalization, and death, as a function of time elapsed since the first dose.

**Results:** For the Pfizer two-dose regimen, vaccine effectiveness in reducing the current risk of Covid-19 ramps to a peak level of 94.9% (95% confidence interval [CI], 94.5 to 95.2) at 2 months (post the first dose) and drops to 70.1% (95% CI, 68.9 to 71.2) after 7 months; effectiveness in reducing the current risk of hospitalization ramps to a peak level of 96.4% (95% CI, 94.7 to 97.5) at 2 months and remains at 87.7% (95% CI, 84.3 to 90.4) at 7 months; effectiveness in reducing the current risk of death ramps to 95.9% (95% CI, 92.9 to 97.6) at 2 months and is maintained at 88.4% (95% CI, 83.0 to 92.1) at 7 months. For the Moderna two-dose regimen, vaccine effectiveness in reducing the current risk of Covid-19 ramps to a peak level of 96.0% (95% CI, 95.6 to 96.4) at 2 months and drops to 81.9% (95% CI, 81.0 to 82.7) after 7 months; effectiveness in reducing the current risk of hospitalization ramps to a peak level of 97.5% (95% CI, 96.3 to 98.3) at 2 months and remains at 92.3% (95% CI, 89.7 to 94.3) at 7 months; effectiveness in reducing the current risk of death ramps to 96.0% (95% CI, 91.9 to 98.0) at 3 months and remains at 93.7% (95% CI, 90.2 to 95.9) at 7 months. For the Janssen one-dose regimen, effectiveness in reducing the current risk of Covid-19 ramps to a peak level of 79.0% (95% CI, 77.1 to 80.7) at 1 month and drops to 64.3% (95% CI, 62.3 to 66.1) after 5 months; effectiveness in reducing the current risk of hospitalization ramps to a peak level of 89.8% (95% CI, 78.8 to 95.1) at 2 months and stays above 80% through 5 months; effectiveness in reducing the current risk of death ramps to 89.4% (95% CI, 52.3 to 97.6) at 3 months and stays above 80% through 5 months. For all three vaccines, the ramping and waning patterns are similar for individuals who were vaccinated at different dates, and across various demographic subgroups (age, sex, race/ethnicity, geographic region, county-level vaccination rate).

**Conclusions:** The two mRNA vaccines are remarkably effective and durable in reducing the risks of hospitalization and death. The Janssen vaccine also offers a high level of protection against hospitalization and death. The Moderna vaccine is significantly more durable than the Pfizer vaccine in reducing the risk of Covid-19. Waning vaccine effectiveness is caused primarily by declining immunity rather than emergence of new variants. It would be worthwhile to investigate the effectiveness of the Janssen vaccine as a two-dose regimen, with the second dose given approximately 1-2 months after the first dose.

---

Severe acute respiratory syndrome coronavirus 2 (SARS-CoV-2) has resulted in ∼45 million cases of coronavirus disease 2019 (Covid-19) and ∼750 thousand deaths in the United States alone. One key step in ending this pandemic is the deployment of durably effective vaccines. Between December 2020 and February 2021, three Covid-19 vaccines, BNT162b2 (Pfizer–BioNTech), mRNA-1273 (Moderna), and Ad26.COV2.S (Janssen), received Emergency Use Authorization (EUA) from the Food and Drug Administration (FDA) on the basis of short-term safety and efficacy against Covid-19^1-3^ and were deployed in the country soon after. However, a recent spike of breakthrough infections has raised concerns about the long-term effectiveness of these vaccines and their effectiveness against new variants, such as the Delta variant.

Several observational studies have assessed waning vaccine effectiveness over time.^4-13^ However, the follow-up was typically shorter than 6 months, and some studies had small sample sizes. Estimates of vaccine effectiveness over broad time periods (1-3 months) were reported, and the confidence intervals for different time periods usually overlapped. These estimates are not informative about the level of waning at the end of the observation period. Most studies compared people who were vaccinated during different calendar time periods or people who had been vaccinated for different amounts of time but did not simultaneously consider both time indexes, such that it is unclear whether waning effectiveness was due to declining immunity to the primary vaccination or to emergence of new variants that are more highly transmissible.

The surveillance systems in the United States seek to capture the vaccination histories and Covid-19 disease outcomes for all the residents and provide valuable resources to assess the effectiveness of Covid-19 vaccines in a real-world setting. Here we report estimates of the effectiveness of the three vaccines currently employed in the United States in reducing the occurrences of symptomatic Covid-19 disease, hospitalization, and death for a period of up to 9 months after vaccination, using the surveillance data from the entire state of North Carolina, which has a population of ∼10.6 million people.

## METHODS

### Data Sources

The state of North Carolina collects data from several sources and partners to monitor the COVID-19 pandemic in the state. The following data sources were used in our study.

#### North Carolina COVID-19 Surveillance System (NC COVID)

NC COVID is a web-based central repository of person-based public health laboratory and communicable disease investigation database used by the North Carolina Department of Health and Human Services, Division of Public Health (DPH), and the state’s 86 local and multi-county district health departments. Laboratories report Covid-19 test results electronically to NC COVID, which initiate investigation workflows for results meeting the public health case definition. Local health jurisdictions collect and enter additional data on known Covid-19 cases, deaths, and demographic information during routine case investigation activities. NC COVID is a component of the Centers for Disease Control and Prevention (CDC) National Notifiable Disease Surveillance System (NNDSS).

#### COVID-19 Vaccine Management System (CVMS)

North Carolina’s information on people vaccinated comes from the COVID-19 Vaccine Management System (CVMS), a secure, cloud-based system that enables vaccine management and data sharing across recipients, care providers, hospitals, agencies, and local, state, and federal governments on one common platform. The system tracks information on provider enrollment, vaccine products administered to individuals, schedules appointments based on the recommended vaccination schedule, and allows the state to manage vaccine supply.

#### Population Census

We used the 2020 Bridged-Race Population estimates produced by the US Census Bureau in collaboration with the National Center for Health Statistics (NCHS) for demographic populations.

#### Analysis Dataset

We extracted data on vaccination histories and clinical outcomes (Covid-19, hospitalization, death) for individuals vaccinated in North Carolina by a North Carolina state provider or federal pharmacy provider during the period from December 13, 2020 through September 8, 2021 by linking data from the NC COVID and CVMS systems (Supplementary Methods). We used the Esri 2020 NC Zip Code population to determine the total number of residents with each combination of demographic variables (age, sex, race/ethnicity, geographic region, county-level vaccination rate).

### Statistical Analysis

The effectiveness of a vaccine at a given time may depend on the vaccination cohort (i.e., people who are vaccinated at a particular calendar date or calendar period) because of changing viral transmission rates over calendar time, as well as on the time elapsed since vaccination because of ramping and declining immunity over that time index. The vaccine effectiveness on the disease incidence over a certain time period (e.g., 60-90 days after the first dose) for individuals who were vaccinated on a particular date (e.g., February 1, 2021) is the unity minus the ratio of the cumulative incidence over that time period for the individuals who were vaccinated on that particular date to that of the unvaccinated individuals. The Kaplan-Meier method^14^ is used to estimate the cumulative incidence of disease over a certain time period for individuals who were vaccinated on a particular day and for those who were not vaccinated. The corresponding estimates of period-specific vaccine effectiveness are averaged over consecutive calendar dates (e.g., February 1–March 31, 2021) to obtain an estimate of the period-specific vaccine effectiveness for that vaccination cohort.

The Cox regression model^15-16^ is used to evaluate vaccine effectiveness in reducing the hazard rate or current risk of disease over time while adjusting for confounding factors. Time to disease is measured from a common calendar date,^16^ namely December 13, 2020, for all individuals in order to control time-varying confounders (e.g., levels of community transmission, prevalence of the Delta variant) by comparing disease incidence between vaccinated and unvaccinated individuals at the same calendar day. The effect of a vaccine on the current risk of disease depends on the time elapsed since the first dose.^16^ This time-varying effect is characterized by a piecewise linear function of time elapsed since the first dose for the log hazard ratio,^16^ with a change-point placed at nearly every month. The effects of the one-dose Pfizer vaccine, two-dose Pfizer vaccine, one-dose Moderna vaccine, two-dose Moderna vaccine, and Janssen vaccine are estimated under the same model. Demographic variables (i.e., age, sex, race/ethnicity, geographic region, county-level vaccination rate) are included as covariates to adjust for potential confounding by individual characteristics and geographic location. The time-varying log hazard ratios for the vaccine effects are estimated by maximum partial likelihood. The vaccine effectiveness on the current risk of disease is estimated by the unity minus the estimated hazard ratio; corresponding 95% confidence intervals are constructed.

## RESULTS

### Study population

Table 1 summarizes the demographic characteristics of the North Carolina population, together with the vaccine uptakes and clinical outcomes in the state during the period of December 13, 2020 – September 8, 2021. North Carolina has a diverse population that reflects the age and sex distributions of the United States, though it has a higher percentage of Black or African Americans and lower percentages of Hispanic or Latino and Asian Americans than the national averages.

**Table 1.** Population characteristics, vaccine uptakes, and clinical outcomes

| Characteristic | Pop. | Vaccine uptakes |  |  | Clinical outcomes |  |  |
| --- | --- | --- | --- | --- | --- | --- | --- |
|  | count | Pfizer | Moderna | Janssen | COVID | Hosp. | Death |
| **All** | 10,600,823 | 3,332,258 (31%) | 2,329,361 (22%) | 345,848 (3%) | 812,494 (8%) | 20,232 | 7,461 |
| **Age** |  |  |  |  |  |  |  |
| <18 | 2,306,400 | 391,651 (17%) | 822 (0%) | 212 (0%) | 137,922 (6%) | 377 | 5 |
| 18 – 34 | 2,432,304 | 747,506 (31%) | 437,366 (18%) | 103,295 (4%) | 246,853 (10%) | 1,281 | 85 |
| 35 – 49 | 1,991,144 | 734,324 (37%) | 490,411 (25%) | 89,947 (5%) | 175,733 (9%) | 2,484 | 379 |
| 50 – 64 | 2,056,433 | 774,368 (38%) | 651,678 (32%) | 109,396 (5%) | 152,042 (7%) | 5,343 | 1,362 |
| 65+ | 1,814,542 | 684,409 (38%) | 749,084 (41%) | 42,998 (2%) | 99,944 (6%) | 10,747 | 5,630 |
| **Sex** |  |  |  |  |  |  |  |
| Female | 5,448,241 | 1,841,400 (34%) | 1,286,325 (24%) | 159,523 (3%) | 430,927 (8%) | 9,960 | 3,460 |
| Male | 5,152,582 | 1,490,858 (29%) | 1,043,036 (20%) | 186,325 (4%) | 381,567 (7%) | 10,272 | 4,001 |
| **Race** |  |  |  |  |  |  |  |
| American Indian or Alaska Native | 180,238 | 35,995 (20%) | 27,518 (15%) | 3,826 (2%) | 15,764 (9%) | 360 | 121 |
| Asian or Pacific Islander | 391,163 | 194,927 (50%) | 86,399 (22%) | 15,330 (4%) | 21,820 (6%) | 267 | 104 |
| Black or African American | 2,453,861 | 720,832 (29%) | 515,991 (21%) | 66,998 (3%) | 200,897 (8%) | 5,707 | 1,833 |
| White | 7,575,561 | 2,380,504 (31%) | 1,699,453 (22%) | 259,694 (3%) | 574,013 (8%) | 13,898 | 5,403 |
| **Ethnicity** |  |  |  |  |  |  |  |
| Hispanic | 1,052,435 | 311,559 (30%) | 159,283 (15%) | 39,248 (4%) | 77,974 (7%) | 1,987 | 549 |
| Non-Hispanic | 9,548,388 | 3,020,699 (32%) | 2,170,078 (23%) | 306,600 (3%) | 734,520 (8%) | 18,245 | 6,912 |
| **Geographic region** |  |  |  |  |  |  |  |
| Coastal | 2,934,844 | 688,349 (23%) | 688,646 (23%) | 84,503 (3%) | 231,402 (8%) | 7,233 | 2,296 |
| Piedmont | 6,501,788 | 2,419,987 (37%) | 1,307,054 (20%) | 210,519 (3%) | 494,582 (8%) | 11,221 | 4,233 |
| Mountain | 1,164,191 | 223,922 (19%) | 333,661 (29%) | 50,826 (4%) | 86,510 (7%) | 1,778 | 932 |
| **County-level vaccination rate** |  |  |  |  |  |  |  |
| <53% | 1,995,109 | 364,744 (18%) | 435,910 (22%) | 43,169 (2%) | 170,101 (9%) | 4,881 | 1,913 |
| 53% – 61% | 1,908,083 | 409,534 (21%) | 500,383 (26%) | 58,197 (3%) | 154,401 (8%) | 6,099 | 1,811 |
| >61% | 6,697,631 | 2,557,980 (38%) | 1,393,068 (21%) | 244,482 (4%) | 487,992 (7%) | 9,252 | 3,737 |
| **Vaccination cohort** |  |  |  |  |  |  |  |
| Dec 13 – Mar 31 | 3,286,055 | 1,626,042 (49%) | 1,537,695 (47%) | 122,318 (4%) | 141,755 (4%) | 3,929 | 1,050 |
| Apr 1 – Jun 30 | 1,889,927 | 1,130,985 (60%) | 575,148 (30%) | 183,794 (10%) | 105,307 (6%) | 2,248 | 70 |
| Jul 1 – Sep 8 | 831,485 | 575,231 (69%) | 216,518 (26%) | 39,736 (5%) | 40,727 (5%) | 935 | 40 |

Vaccination rates are higher among older adults, females, and Whites and Asians or Pacific Islanders. The Pfizer vaccine was the most commonly administered, followed not far behind by the Moderna vaccine. The Janssen vaccine was taken by only a small percentage of the population, and at much later dates than the two mRNA vaccines.

During the period of December 13, 2020 – September 8, 2021, a total of 812,665 cases of Covid-19 were reported, 20,241 of which were known to lead to hospitalization and 7,462 of which were known to lead to death. However, the hospitalization and death status were known for only approximately 40% and 60% of Covid-19 cases, respectively. Thus, the actual numbers of hospitalization and death were substantially higher.

### Vaccine effectiveness on the disease incidence over time by vaccination cohort

For the Moderna and Pfizer vaccines, effectiveness ramps to a peak in the mid 90% range around 2 months post the first dose and then gradually declines. The Moderna vaccine appears more durable than the Pfizer vaccine. For the Janssen vaccine, effectiveness ramps to a peak in the low 70% around 1-2 months post the first dose and declines gradually afterward.

For all three vaccines, especially the two mRNA vaccines, the ramping and waning patterns are similar among vaccination cohorts. For example, the effectiveness of the Moderna vaccine is in the low 90% at months 3-4 for all vaccination cohorts and is around 80% at months 7-8 for all vaccination cohorts. These results suggest that waning vaccine effectiveness was caused mainly by decline of immunity to the primary vaccination, rather than by emergence of new variants that are more highly transmissible. Thus, we assumed in the Cox regression analysis that vaccine effectiveness does not depend on vaccination cohort.

Because the Delta variant accounts for the majority of Covid-19 cases in the state of North Carolina since July, 2021, the effectiveness estimates on or below the bottom-left to the top-right diagonal line of Table 2 pertain mainly to the Delta variants, whereas those above the diagonal line represent more of the older variants. The two sets of estimates for the same amount of time elapsed since vaccination are similar, suggesting that the prevalence of the Delta variant did not have a major impact on vaccine effectiveness.

**Table 2.** Effectiveness of the BNT162b2 (Pfizer-BioNTech) two-dose vaccine, mRNA-1273 (Moderna) two-dose vaccine, and Ad26.COV2.S (Janssen) one-dose vaccine in reducing the incidence of COVID-19 over successive time periods by vaccination cohort

| Vaccination cohort | Months since vaccination |  |  |  |  |  |  |  |  |
| --- | --- | --- | --- | --- | --- | --- | --- | --- | --- |
|  | 0 – 1 | 1 – 2 | 2 – 3 | 3 – 4 | 4 – 5 | 5 – 6 | 6 – 7 | 7 – 8 | 8 – 9 |
| **Pfizer-BioNtech** |  |  |  |  |  |  |  |  |  |
| All dates | 67.2% | 92.4% | 91.0% | 84.3% | 83.1% | 77.6% | 68.0% | 66.8% | 63.2% |
| Dec 15 – Jan 31 | 45.4% | 90.3% | 91.7% | 89.9% | 86.7% | 78.2% | 60.2% | 62.7% | 60.5% |
| Feb 1 – Feb 28 | 57.9% | 91.0% | 93.0% | 88.2% | 82.4% | 73.1% | 74.8% | 81.1% |  |
| Mar 1 – Mar 31 | 56.7% | 93.1% | 92.3% | 85.0% | 76.8% | 76.7% | 79.1% |  |  |
| Apr 1 – Apr 30 | 62.7% | 92.7% | 88.5% | 81.8% | 81.3% | 85.3% |  |  |  |
| May 1 – May 31 | 68.0% | 91.5% | 88.6% | 88.0% | 90.0% |  |  |  |  |
| June 1 – June 30 | 75.0% | 94.0% | 91.1% | 87.1% |  |  |  |  |  |
| July 1 – July 31 | 84.3% | 94.3% | 90.6% |  |  |  |  |  |  |
| Aug 1 – Sept 8 | 93.5% | 96.3% |  |  |  |  |  |  |  |
| **Moderna US** |  |  |  |  |  |  |  |  |  |
| All dates | 69.4% | 91.6% | 93.5% | 92.3% | 89.2% | 85.8% | 82.6% | 80.8% | 85.1% |
| Dec 15 – Jan 31 | 38.4% | 82.7% | 92.3% | 91.6% | 88.5% | 86.0% | 81.3% | 80.4% | 84.0% |
| Feb 1 – Feb 28 | 62.6% | 90.6% | 90.0% | 94.8% | 90.9% | 83.8% | 82.4% | 78.9% |  |
| Mar 1 – Mar 31 | 71.3% | 94.0% | 95.5% | 91.7% | 87.2% | 85.0% | 85.9% |  |  |
| Apr 1 – Apr 30 | 71.6% | 92.9% | 94.5% | 90.7% | 89.1% | 90.5% |  |  |  |
| May 1 – May 31 | 68.8% | 94.2% | 93.2% | 92.3% | 91.6% |  |  |  |  |
| June 1 – June 30 | 78.1% | 94.2% | 94.9% | 93.6% |  |  |  |  |  |
| July 1 – July 31 | 86.3% | 96.0% | 95.8% |  |  |  |  |  |  |
| Aug 1 – Sept 8 | 92.0% | 95.0% |  |  |  |  |  |  |  |
| **Janssen** |  |  |  |  |  |  |  |  |  |
| All dates | 58.1% | 73.4% | 70.3% | 55.2% | 60.8% | 59.8% | 73.5% |  |  |
| Feb 1 – Mar 15 | 64.7% | 81.5% | 78.5% | 79.5% | 58.8% | 60.8% | 73.5% |  |  |
| Mar 16 – Apr 15 | 55.0% | 79.9% | 67.4% | 57.8% | 51.7% | 59.0% |  |  |  |
| Apr 16 – Sept 8 | 58.0% | 70.3% | 69.6% | 45.7% | 73.6% |  |  |  |  |

### Vaccine effectiveness on the current risk of symptomatic Covid-19 over time

Estimates of vaccine effectiveness in reducing the current risk of acquiring symptomatic Covid-19 are displayed in Figure 1A. For the Pfizer two-dose regimen, vaccine effectiveness ramps to a peak level of 94.9% (95% CI, 94.5 to 95.2) at 2 months (post the first dose). Effectiveness starts to decline after 2 months and drops to 70.1% (95% CI, 68.9 to 71.2) after 7 months.

**Figure 1.**
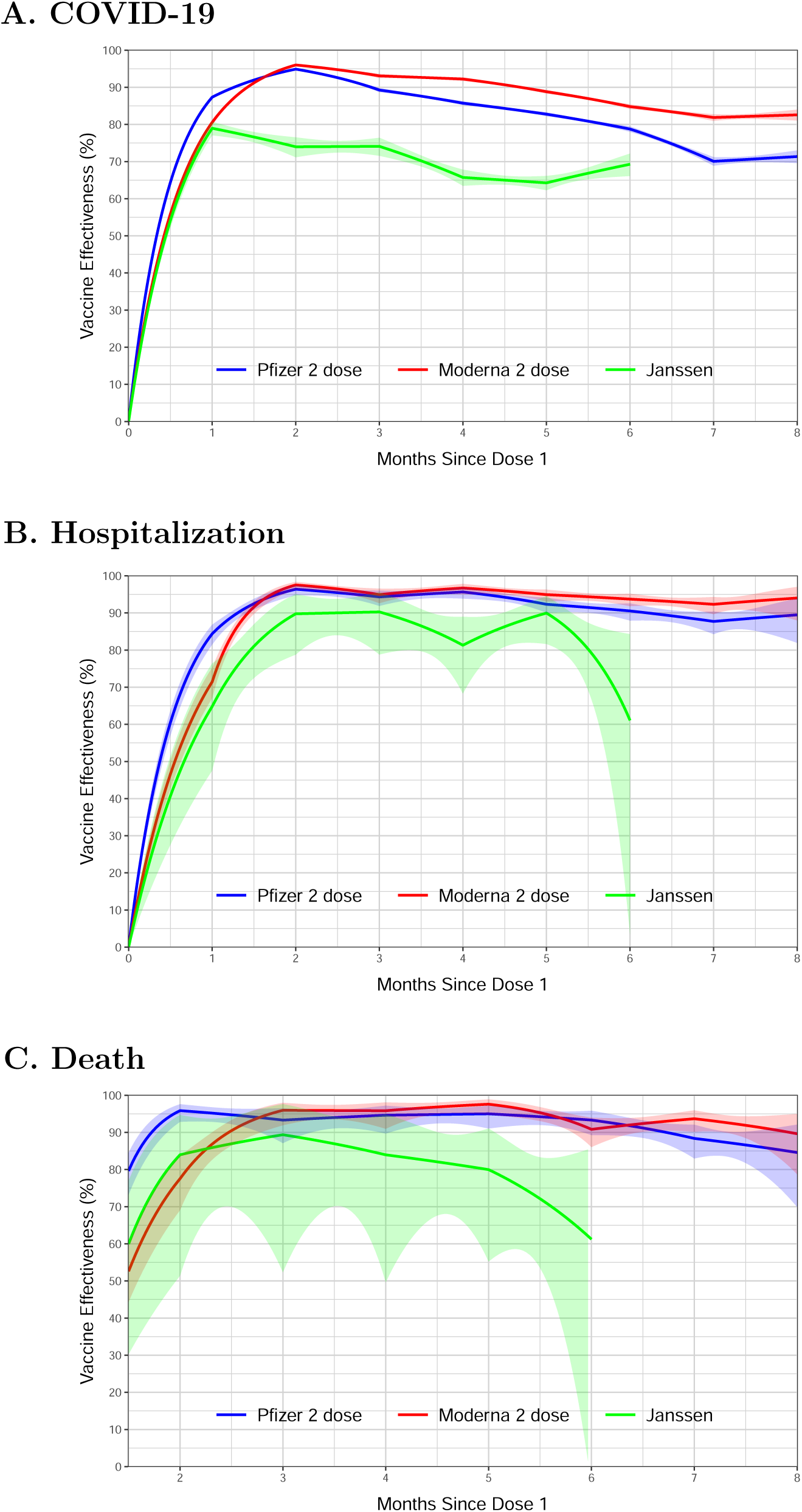
Effectiveness of the BNT162b2 (Pfizer–BioNTech), mRNA-1273 (Mod-erna), and Ad26.COV2.S (Janssen) in reducing the risks of COVID-19 disease (A), hospitalization (B), and death (C) in the state of North Carolina, December 13, 2020 – September 8, 2021. Estimates are shown by solid curves, and 95% confidence intervals are shown by shaded bands.

For the Moderna two-dose regimen, vaccine effectiveness ramps to a peak level of 96.0% (95% CI, 95.6 to 96.4) at 2 months. Effectiveness starts to decline after 2 months. Effectiveness is maintained at 81.9% (95% CI, 81.0 to 82.7) after 7 months.

For the Janssen one-dose regimen, vaccine effectiveness ramps to a peak level of 79.0% (95% CI, 77.1 to 80.7) at 1 month. Effectiveness starts to decline after 1 month and drops to 64.3% (95% CI, 62.3 to 66.1) after 5 months. Because the Janssen vaccine was not deployed until March 2021, the information about its effectiveness beyond 5 months is limited.

For all three vaccines, the ramping and waning patterns are similar across various demographic subgroups (age, sex, race/ethnicity, geographic region, county-level vaccination rate) (Fig. S1-S5). For all three vaccines, effectiveness is lower in the 65+ age group than other age groups (Fig. S1) and is higher in counties with higher vaccination rates (Fig. S5).

### Vaccine effectiveness on the current risk of hospitalization over time

Estimates of vaccine effectiveness in reducing the current risk of acquiring severe Covid-19 that leads to hospitalization are displayed in Figure 1B. For the Pfizer two-dose regimen, vaccine effectiveness ramps to a peak level of 96.4% (95% CI, 94.7 to 97.5) at 2 months and maintains at 87.7% (95% CI, 84.3 to 90.4) at 7 months. For the Moderna two-dose regimen, vaccine effectiveness ramps to a peak level of 97.5% (95% CI, 96.3 to 98.3) at 2 months and remains at 92.3% (95% CI, 89.7 to 94.3) at 7 months. For the Janssen one-dose regimen, vaccine effectiveness ramps to a peak level of 89.8% (95% CI, 78.8 to 95.1) at 2 months and stays above 80% through 5 months. For the two mRNA vaccines, effectiveness is lower in the 65+ age group than other age groups (Fig. S6) and is higher in counties with higher vaccination rates (Fig. S10).

### Vaccine effectiveness on the current risk of death over time

Estimates of vaccine effectiveness in reducing the current risk of death due to Covid-19 are displayed in Figure 1C. For the Pfizer two-dose regimen, vaccine effectiveness ramps to 95.9% (95% CI, 92.9 to 97.6) at 2 months and remains at 88.4% (95% CI, 83.0 to 92.1) at 7 months. For the Moderna two-dose regimen, effectiveness ramps to 96.0% (95% CI, 91.9 to 98.0) at 3 months and remains at 93.7% (95% CI, 90.2 to 95.9) at 7 months. For the Janssen one-dose regimen, vaccine effectiveness ramps to 89.4% (95% CI, 52.3 to 97.6) at 3 months and stays above 80% through 5 months. Effectiveness is lower in the 65+ age group than the 18-64 age group (Fig. S11).

## DISCUSSION

The estimates of vaccine effectiveness from this study are consistent with and complement the estimates of vaccine efficacy from phase 3 trials.^1-3,17-18^ Specifically, the peak levels for the three vaccines are similar between this study and phase 3 trials, although phase 3 trials were not powered to determine when the peak occurs. The large sample size not only allowed us to pinpoint the location of the peak but also to estimate vaccine effectiveness against hospitalization and death over time. The long-term vaccine effectiveness on symptomatic Covid-19 reported here is lower than what was shown by limited phase 3 data,^17-18^ suggesting that behavioral changes (in face masking, social distancing, and other mitigation measures) might dilute the biological effects of vaccines in the real world.

Our results show that the effectiveness of the two mRNA vaccines is remarkably high and durable against hospitalization and death and that the Moderna vaccine has significantly longer duration of protection than the Pfizer vaccine, particularly against symptomatic Covid-19. In addition, the Janssen vaccine offers a high level of protection against hospitalization and death. Thus, the unvaccinated people in the United States should be strongly encouraged to get vaccinated.

Our results also show that the effectiveness of the Janssen vaccine ramps to a peak level similar to that of the two mRNA vaccines one month after vaccination and then starts to decline afterward. Thus, it would be worthwhile to investigate the use of the Janssen vaccine as a two-dose regimen given approximately 1-2 months apart. However, a one-dose regimen is useful for transient populations and developing countries. Thus, the high effectiveness of the one-dose regimen seen in our data is encouraging.

Our study is observational in nature and thus suffers from confounding bias. We have adjusted for measured confounders (age, sex, race/ethnicity, geographical region, and county-level vaccination rate). More important, we measured time to disease occurrence from the start of the study in order to compare disease incidence between the vaccinated and unvaccinated individuals at the same calendar time, thus avoiding confounding due to time trends (e.g., level of community transmission, prevalence of new variants). However, the people who choose not to be vaccinated may differ from those who do, even after adjusting for all demographic variables (age, sex, race/ethnicity, geographic region, county-level vaccination rate), and the differences may become more appreciable over time. In addition, individuals with Covid-19 symptoms were unlikely to be vaccinated, and some “unvaccinated” individuals might have been vaccinated outside of primary healthcare systems. While acknowledging that there is likely some unknown residual confounding bias, we believe that our estimates represent vaccine effectiveness in the real world, which is more relevant to public health than the biological effects which we cannot reliably estimate.

There are high proportions of missing data on hospitalization and death, in part due to limited availability of local case investigators to investigate disease outcomes. Because special effort has been made to specifically request reporting of post-vaccination cases resulting in hospitalization or death, records on hospitalization and death were less likely to be missing in vaccinated people than in unvaccinated people. This differential missingness by itself would lead to underestimation of vaccine effectiveness against hospitalization and death.

This study included vaccination histories for individuals vaccinated in North Carolina by a North Carolina state provider or federal pharmacy provider.

Recipient-level vaccination data for residents of North Carolina who were vaccinated outside of North Carolina, and for residents vaccinated through a federal entity channel (Department of Defense, Veterans Health Administration, Indian Health Service, and federal Bureau of Prisons) are not available through CVMS and therefore were not included in the analysis. Those data represent an estimated <5% of total vaccine administrations in North Carolina.

Routinely linked laboratory and vaccine registry data from comprehensive statewide surveillance systems are useful for studying vaccine effectiveness. The methodology used in this study^16^ can be applied to surveillance linked laboratory and vaccine data from other states. By combining data from multiple states, we will be able to gain more comprehensive understandings of vaccine effectiveness in the United States, especially in various subgroups and with respect to hospitalization and death. With additional follow-up data, we would be able to not only evaluate the effectiveness of the original vaccine series beyond 9 months but also the effectiveness of the booster programs and the need for additional boosters.

## Data Availability

All data produced in the present study are available upon reasonable request to the North Carolina Department of Public Health

## Supplementary Appendix for

### SUPPLEMENTARY METHODS

#### Data Linkage

Data from the state’s COVID-19 vaccination management system include a person-level view of all doses received from state-affiliated providers. Data from federal pharmacy providers are provided at the dose level. Individuals are free to participate in both vaccination programs may be represented in both views. These data are refreshed weekly.

Records from the state’s lab and case surveillance system are matched to vaccination data weekly. To capture post vaccination cases, positive lab results collected after December 11, 2020 - the date of first vaccination in North Carolina – are selected for matching to vaccination data. Lab records contain name elements, date of birth, and geographic information for linkage purposes, demographic covariates, and COVID-19 outcomes including hospitalization and death.

#### Matching Model

Data from the vaccination registry are transformed prior to matching. State administered records are already normalized at the person level. A person-normalized view of the federal pharmacy data is generated by probabilistically deduplicating dose-level data on name elements, date of birth, and geography prior to matching using Link Plus 3.0^i^ (CDC).

Positive lab results are then matched to person-level records from the state and federal pharmacy vaccine registry across two linkage models. Records in each are probabilistically linked on name elements, dates of birth, and geographic territory in Link Plus, generating a scored list of potential record matches. Results of matching models above a minimum score threshold are selected for manual review.

After manual validation, matching records are added to a cumulative linkage table, capturing record IDs from each system and the date of the linkage. The matching process is continuous; IDs not previously matched are selected in subsequent rounds of linkage. Cumulative linkage tables are used to join the lab and vaccination data to summarize vaccination outcomes.

**Figure S1.**
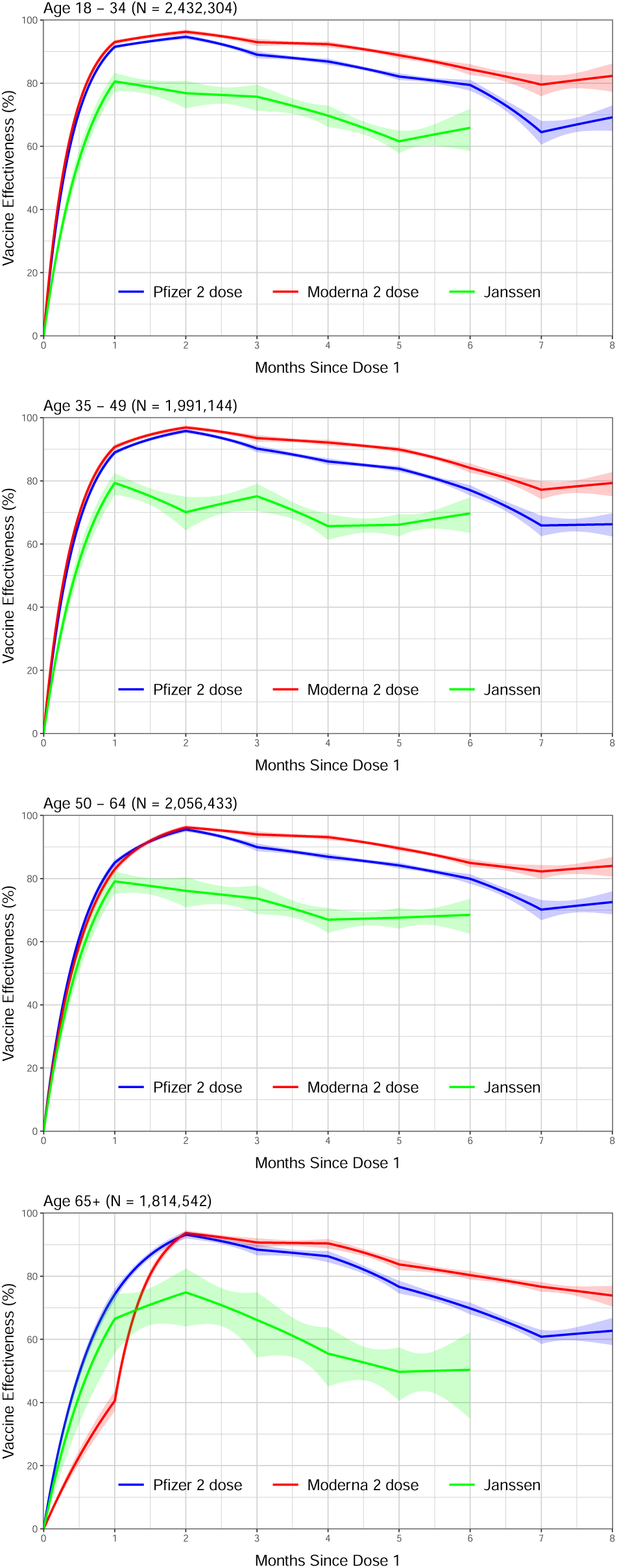
Vaccine effectiveness in reducing the risk of COVID-19 disease in the state of North Carolina, December 13, 2020 – September 8, 2021, by age group. Estimates are shown by solid curves, and 95% confidence intervals are shown by shaded bands.

**Figure S2.**
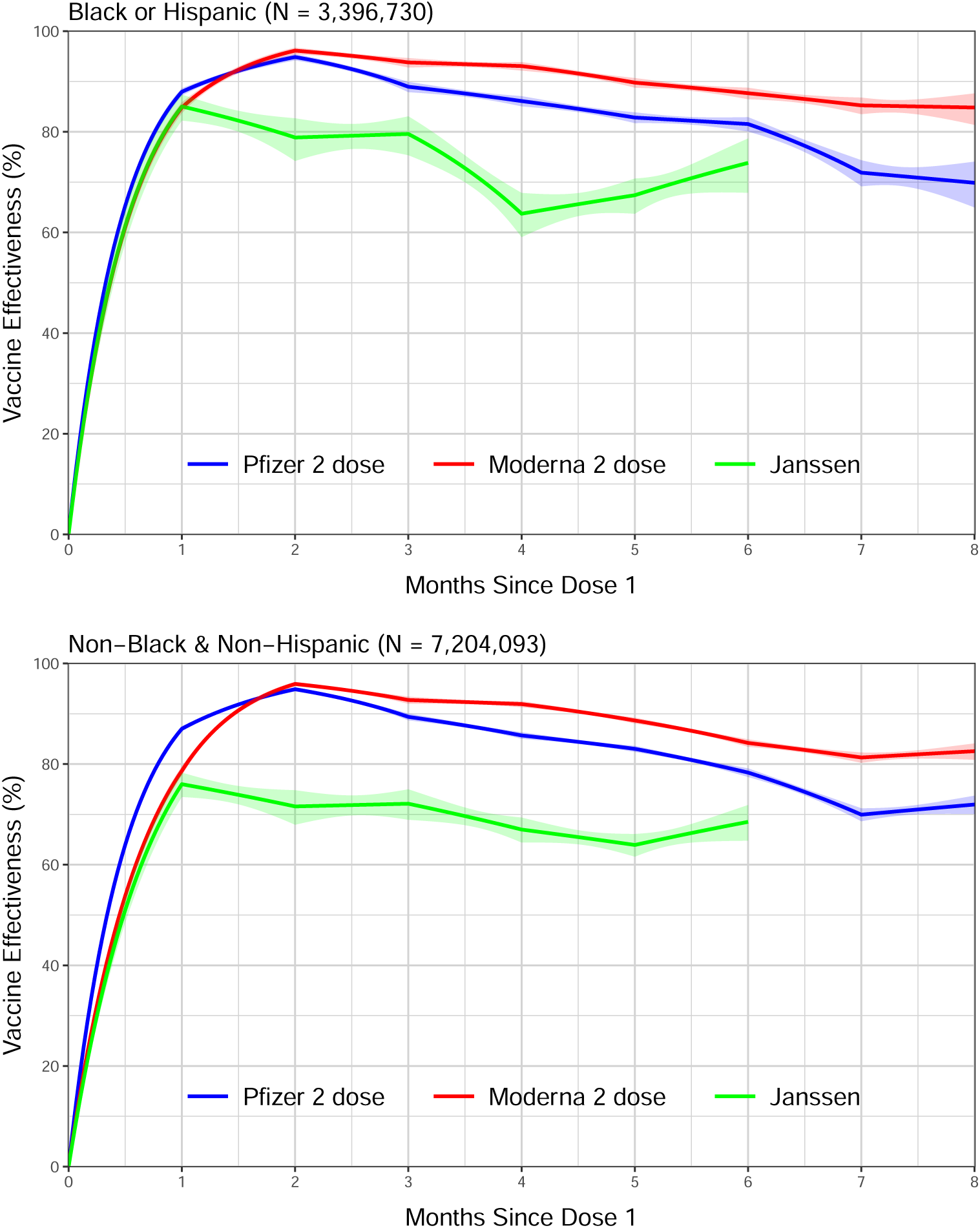
Vaccine effectiveness in reducing the risk of COVID-19 disease in the state of North Carolina, December 13, 2020 – September 8, 2021, by race/ethnicity. Estimates are shown by solid curves, and 95% confidence intervals are shown by shaded bands.

**Figure S3.**
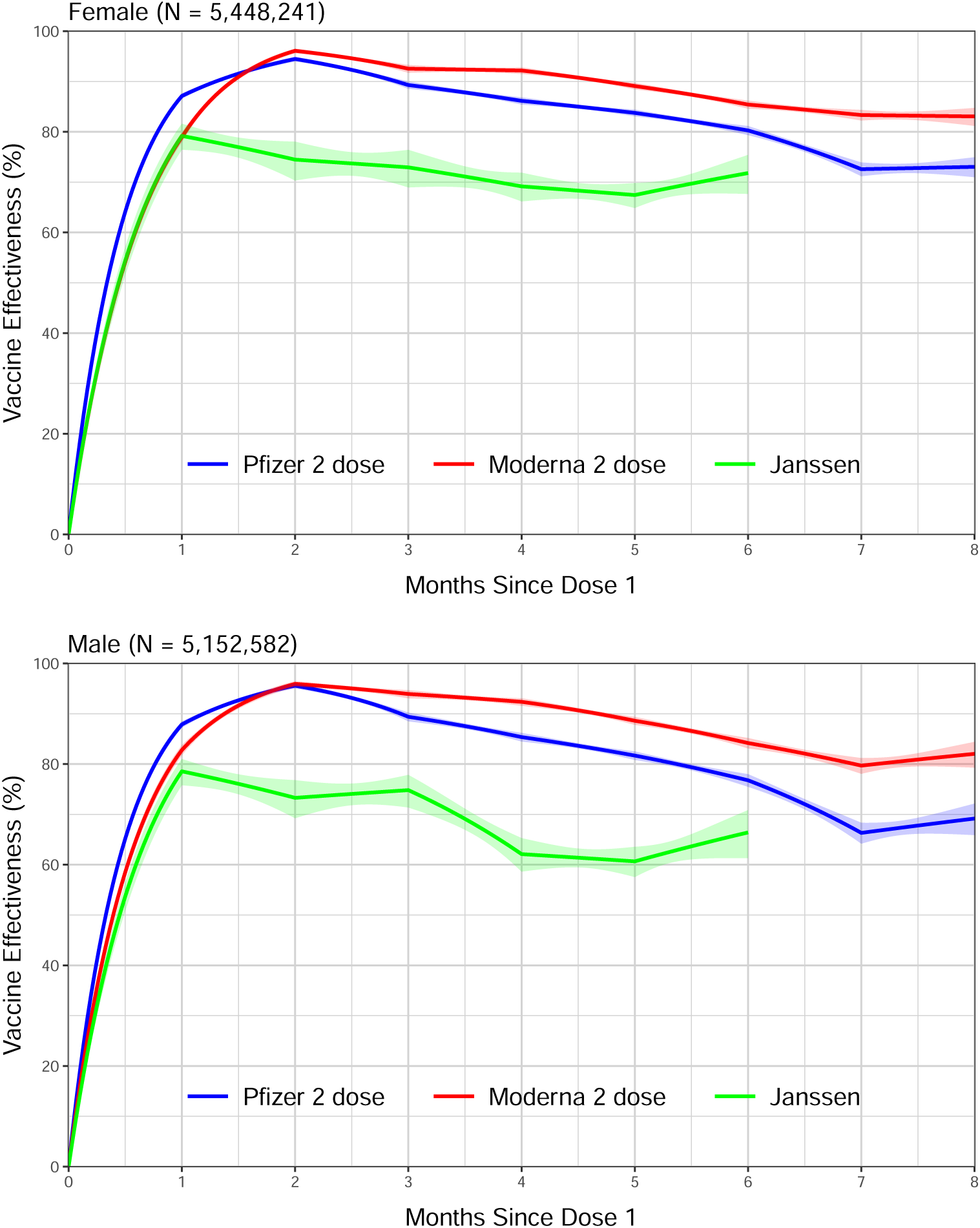
Vaccine effectiveness in reducing the risk of COVID-19 disease in the state of North Carolina, December 13, 2020 – September 8, 2021, by sex. Estimates are shown by solid curves, and 95% confidence intervals are shown by shaded bands.

**Figure S4.**
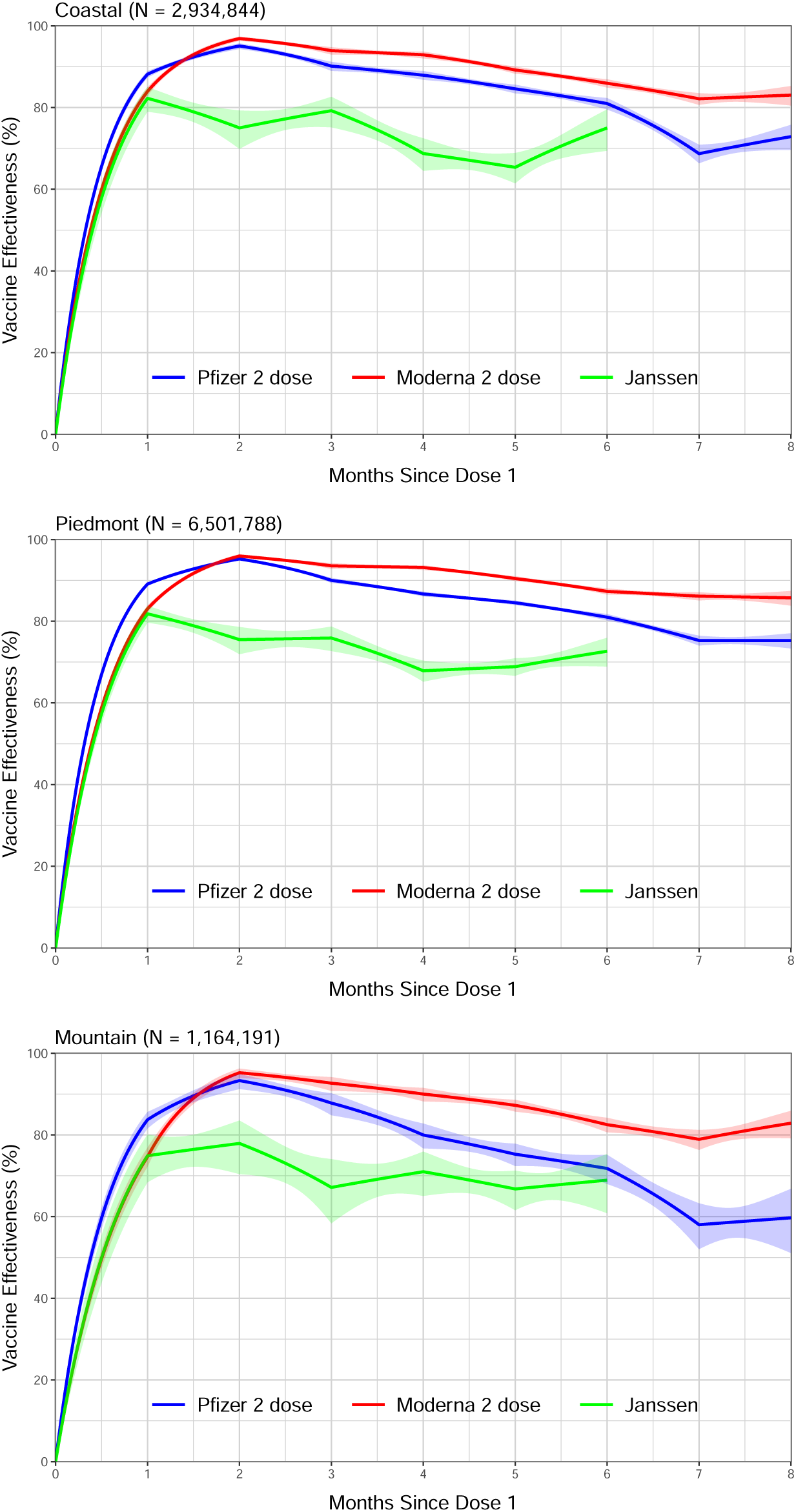
Vaccine effectiveness in reducing the risk of COVID-19 disease in the state of North Carolina, December 13, 2020 – September 8, 2021, by geographic region. Estimates are shown by solid curves, and 95% confidence intervals are shown by shaded bands.

**Figure S5.**
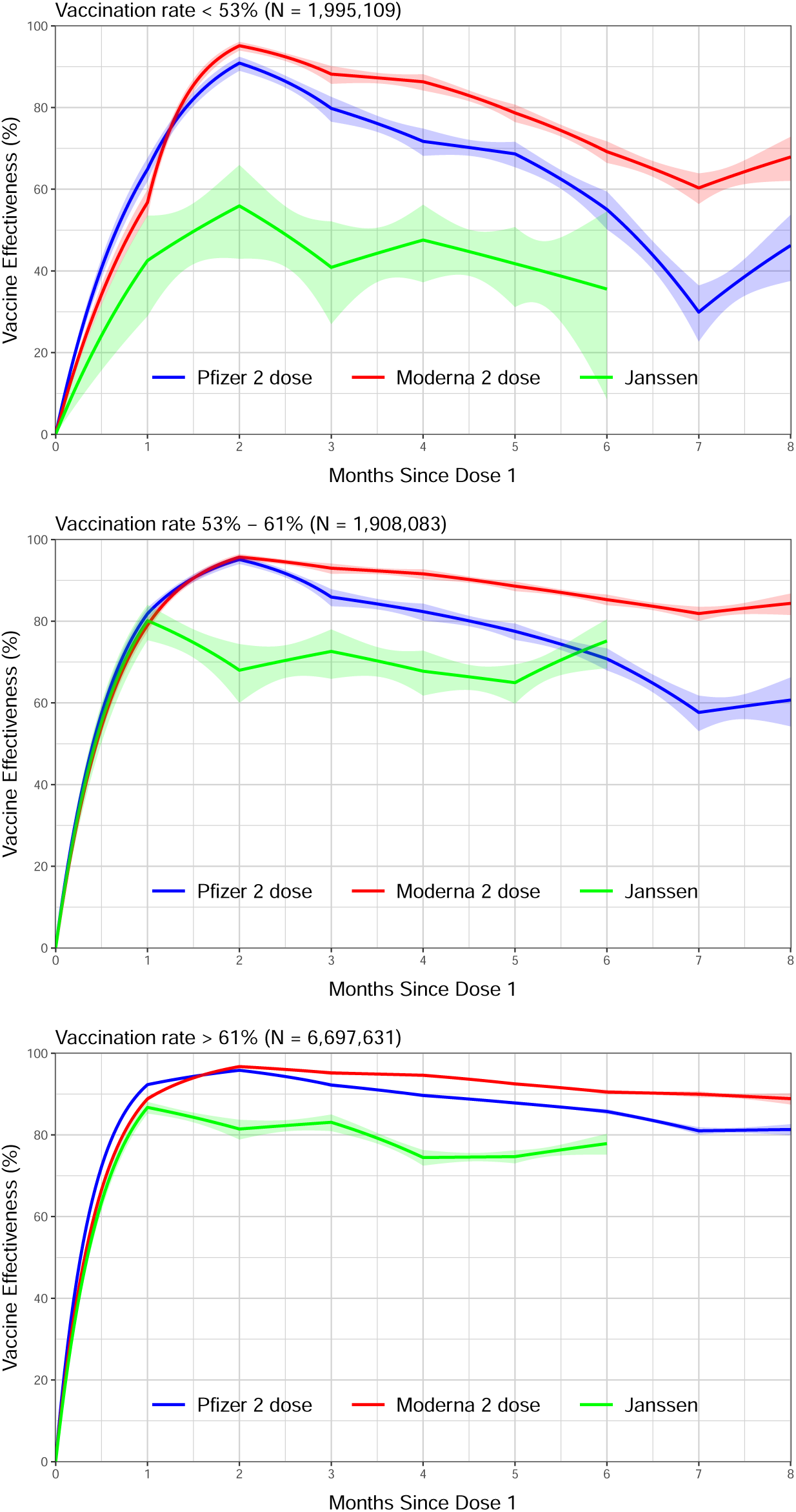
Vaccine effectiveness in reducing the risk of COVID-19 disease in the state of North Carolina, December 13, 2020 – September 8, 2021, by county-level vaccination rate. Estimates are shown by solid curves, and 95% confidence intervals are shown by shaded bands.

**Figure S6.**
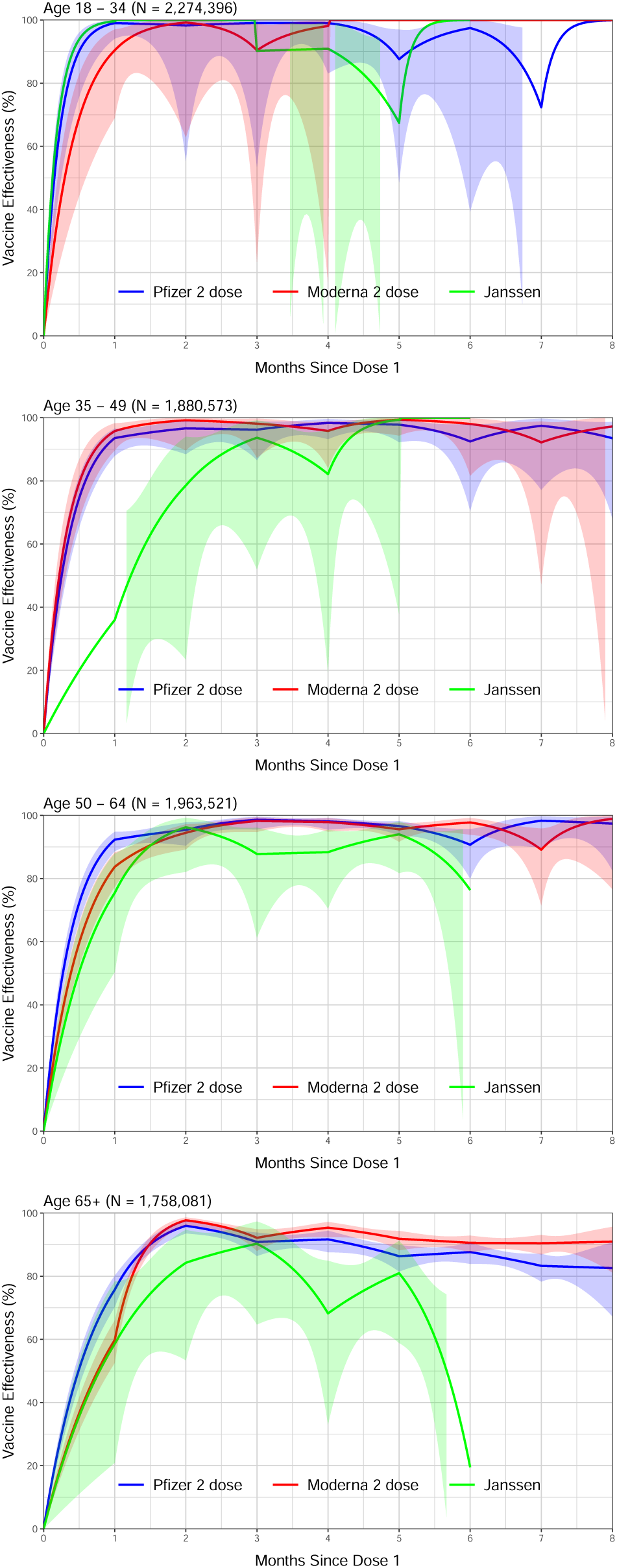
Vaccine effectiveness in reducing the risk of hospitalization in the state of North Carolina, December 13, 2020 – September 8, 2021, by age group. Estimates are shown by solid curves, and 95% confidence intervals are shown by shaded bands.

**Figure S7.**
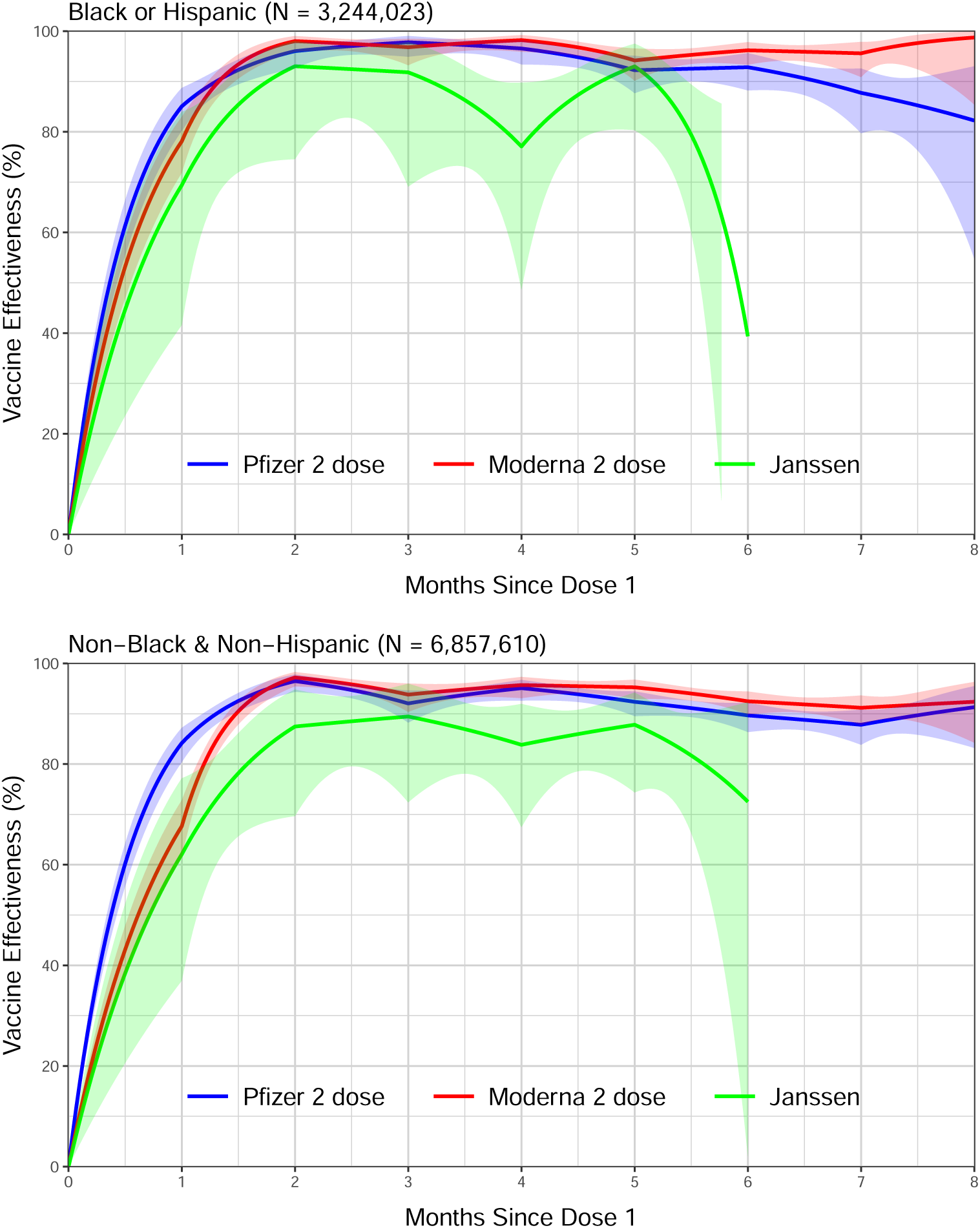
Vaccine effectiveness in reducing the risk of hospitalization in the state of North Carolina, December 13, 2020 – September 8, 2021, by race/ethnicity. Estimates are shown by solid curves, and 95% confidence intervals are shown by shaded bands.

**Figure S8.**
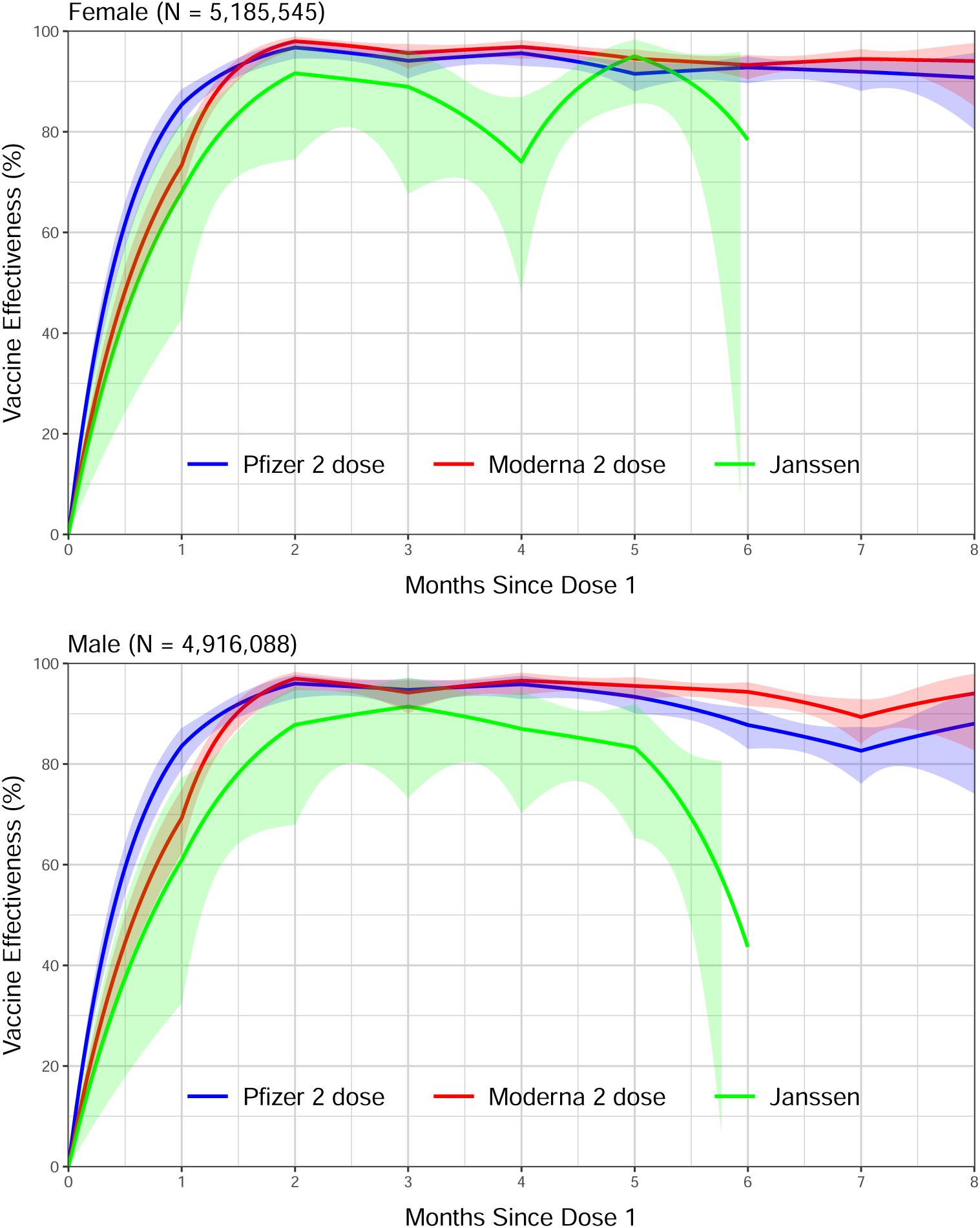
Vaccine effectiveness in reducing the risk of hospitalization in the state of North Carolina, December 13, 2020 – September 8, 2021, by sex. Estimates are shown by solid curves, and 95% confidence intervals are shown by shaded bands.

**Figure S9.**
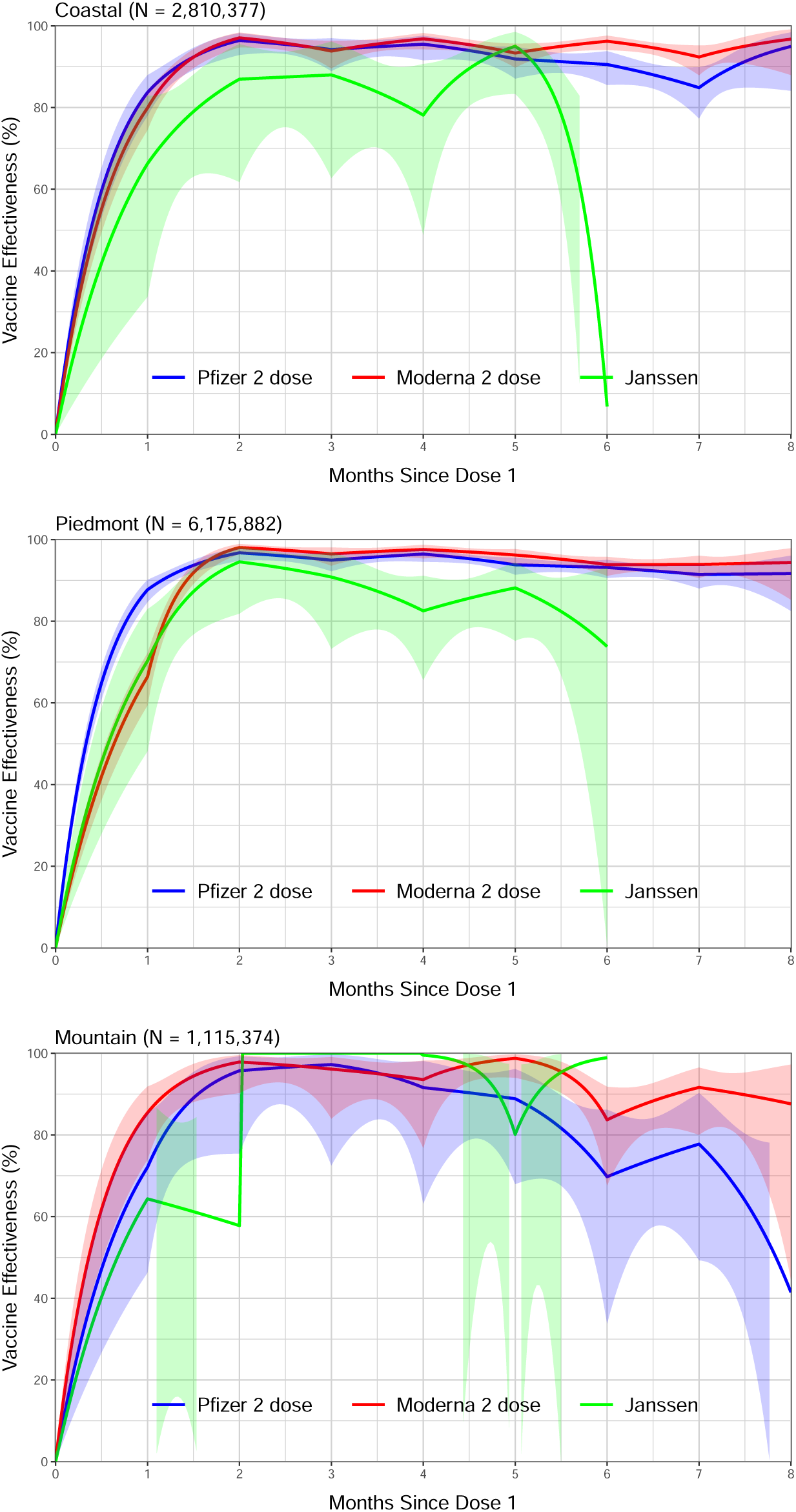
Vaccine effectiveness in reducing the risk of hospitalization in the state of North Carolina, December 13, 2020 – September 8, 2021, by geographic region. Estimates are shown by solid curves, and 95% confidence intervals are shown by shaded bands.

**Figure S10.**
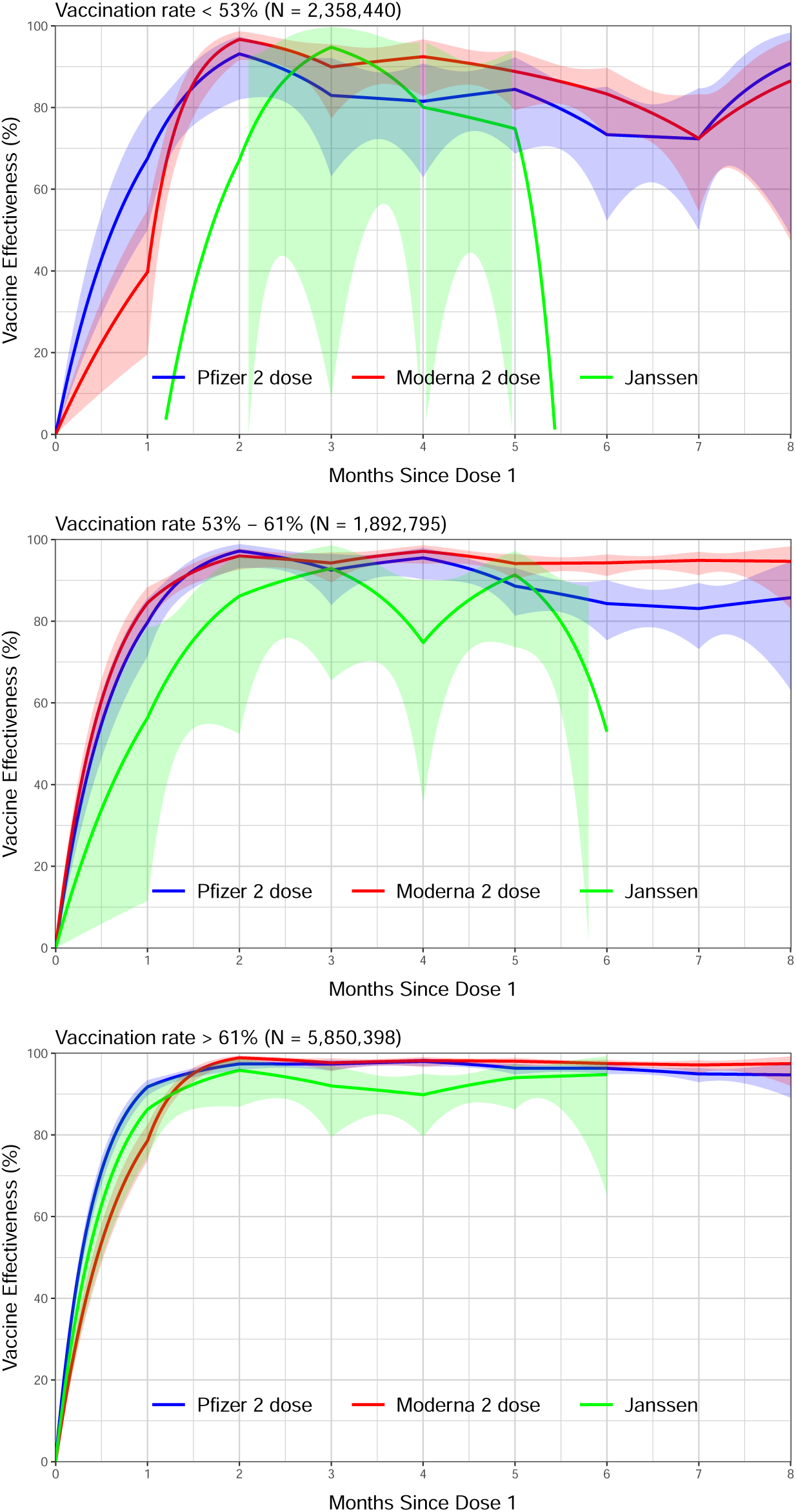
Vaccine effectiveness in reducing the risk of hospitalization in the state of North Carolina, December 13, 2020 – September 8, 2021, by county-level vaccination rate. Estimates are shown by solid curves, and 95% confidence intervals are shown by shaded bands.

**Figure S11.**
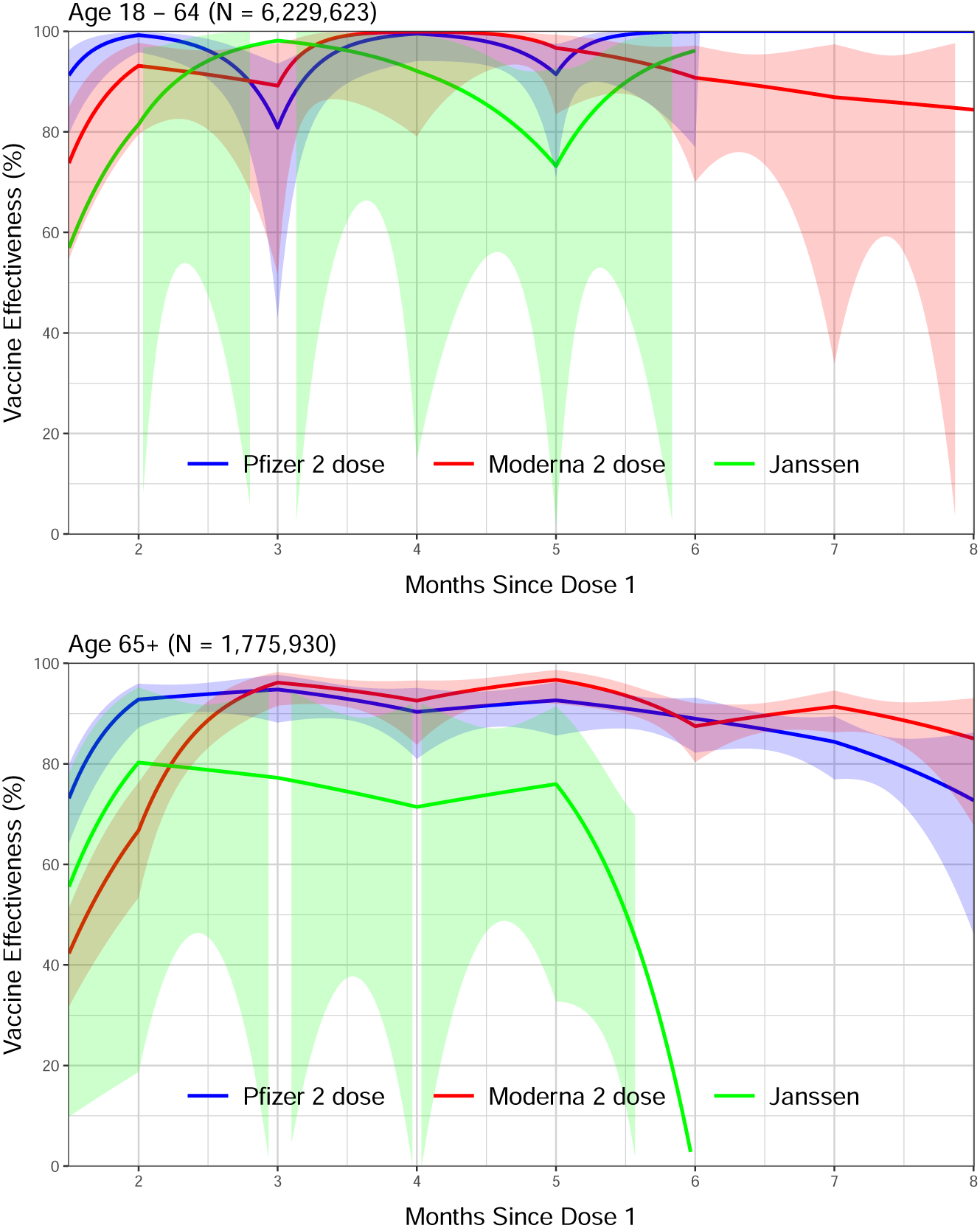
Vaccine effectiveness in reducing the risk of death in the state of North Carolina, December 13, 2020 – September 8, 2021, by age group. Estimates are shown by solid curves, and 95% confidence intervals are shown by shaded bands.

## Footnotes

i Centers for Disease Control and Prevention, National Center for Chronic Disease Prevention and Health Promotion; 2020. Available at: https://www.cdc.gov/cancer/npcr/ https://www.cdc.gov/cancer/npcr/tools/registryplus/lp.htm.

